# Effect of Acipimox on skeletal muscle biochemistry, structure and function in older people with probable sarcopenia: an experimental medicine study

**DOI:** 10.1101/2024.11.28.24318143

**Authors:** Claire McDonald, Craig Alderson, Matthew G Birkbeck, Silvia Del Din, Grainne G Gorman, Kieren G Hollingsworth, Cameron Kirk, Clare Massarella, Lynn Rochester, Helen AL Tuppen, Charlotte Warren, Avan A Sayer, Miles D Witham

**Affiliations:** AGE Research Group, Translational and Clinical Research Institute, Faculty of Medical Sciences, Newcastle University, Newcastle upon Tyne, UK; NIHR Newcastle Biomedical Research Centre, Newcastle upon Tyne Hospitals NHS Foundation Trust, Cumbria, Northumberland, Tyne and Wear NHS Foundation Trust and Newcastle University, Newcastle upon Tyne, UK; Gateshead Health NHS Foundation Trust, Gateshead, UK; Newcastle Upon Tyne Hospitals NHS Foundation Trust, Newcastle Upon Tyne, UK; Newcastle Magnetic Resonance Centre, Translational and Clinical Research Institute, Newcastle University, Newcastle upon Tyne, Tyne and Wear, UK; Wellcome Centre for Mitochondrial Research, Translational and Clinical Research Institute, Newcastle University, Newcastle upon Tyne, Tyne and Wear, UK; Brain and Movement Research Group, Translational and Clinical Research Institute, Newcastle University, Newcastle upon Tyne, Tyne and Wear, UK

**Keywords:** Sarcopenia, acipimox, nicotinamide adenine dinucleotide, clinical trial

## Abstract

**Background:** Skeletal muscle Nicotinamide Adenine Dinucleotide (NAD) concentrations are low in people with sarcopenia. Increasing NAD concentrations may offer a novel therapy. This study tested if Acipimox (a NAD precursor) improves skeletal muscle NAD concentration and function in people with probable sarcopenia.

**Methods:** Participants aged 65 and over with low walk speed (<0.8m/s) and low muscle strength (by 2019 European Working Group criteria) were recruited to this before and after, proof-of-concept study. Participants received acipimox 250mg orally (twice or thrice daily according to creatinine clearance) + aspirin 75mg daily (to prevent facial flushing) for 4 weeks.

Muscle biopsy of the vastus lateralis, ^31^P magnetic resonance spectroscopy, and a 7-digital mobility assessment were performed before starting acipimox and after 3 weeks of treatment. The primary outcome was change in skeletal muscle NAD concentration. Secondary outcomes included change in phosphocreatine recovery rate and measures of physical performance.

**Results:** Eleven participants (8 women), mean age 78.9 years (SD 4.3), were recruited. Mean walk speed at baseline was 0.69m/s (SD 0.07). All completed baseline and follow-up visits. Median medication adherence was 95% (range 91–104%). There was no statistically significant difference in the primary outcome of change in NAD concentrations in skeletal muscle between baseline and follow-up [median difference: −0.003umol/g (IQR −0.058 to 0.210); P=0.26] or secondary outcomes. Nineteen adverse events were reported, none serious.

**Conclusions:** Although the study protocol was feasible and well tolerated, acipimox did not improve skeletal muscle NAD concentration, biochemical markers or physical function in people with probable sarcopenia.

## Background

Sarcopenia is the age-related loss of muscle strength and mass. [1] It is a major cause of adverse health outcomes, including falls, impaired activities of daily living, decreased quality of life, admission to long-term care and early death. [1–3] The only intervention to treat sarcopenia that has unequivocal evidence of efficacy is resistance exercise. [4] However, many older adults are unable or unwilling to undertake resistance exercise or to sustain the intensity of training required for optimal effects. [5] Given the lack of other effective treatments, pharmacological therapies are needed as alternatives to exercise training and to augment its effects.

Nicotinamide Adenine Dinucleotide (NAD) is a key component in cellular energy metabolism. Low skeletal muscle NAD concentrations are found in patients with sarcopenia; skeletal muscle NAD concentrations correlate with grip strength and gait speed. [6]. Preclinical studies in young and aged mice and a murine model of mitochondrial myopathy have suggested that NAD supplementation may increase mitochondrial biogenesis, improve mitochondrial function and increase muscle mass and strength. [7, 8]. In *Caenorhabditis elegans* nematode worms, NAD supplementation increased spontaneous mobility measured using movement trackers. [7] Despite the promise of pre-clinical models, there has been limited progress in translating NAD precursor supplementation into human studies of muscle weakness. NAD precursors have been shown to alter the NAD metabolome but did not directly increase skeletal muscle NAD concentrations. [8] In young athletes, NAD supplementation failed to alter skeletal muscle metabolism, training response or performance. [8] In older adults, results have been mixed [9, 10] with some, but not all, studies indicating that NAD supplementation may improve walk speed [11] and lower-limb strength [10].

Acipimox is a nicotinic acid analogue licenced to treat hyperlipidaemia. Besides the known effects of Acipimox on lipolysis, it has been shown to improve ex-vivo mitochondrial respiration in patients with type 2 diabetes. [12]. Acipimox has a well-established safety record and is generally well-tolerated. The most commonly reported side effects are headache, dyspepsia, and flushing, which can be reduced by co-prescribing low-dose aspirin [13]. Previous clinical trials of NAD precursors, including Acipimox, have not been conducted in patients with sarcopenia – the group with the most to gain from such an intervention and arguably the group most likely to respond to such an intervention.

Therefore, this experimental medicine feasibility trial tested whether Acipimox improved skeletal muscle NAD concentrations, mitochondrial function and physical performance in older people with probable sarcopenia.

## Methods

The study was an open-label, uncontrolled, before-and-after experimental medicine feasibility trial. The protocol has been published previously [14]. The trial was approved by the UK Health Research Authority Northeast - Tyne & Wear South Research Ethics Committee (approval number 21/NE/0100). The trial was also approved by the UK Medicines and Healthcare Products Regulatory Agency (MHRA; EudraCT trial reference number 2021-000993-28). The trial has been included in the National Institute for Health Research Clinical Research Network (NIHR CRN) portfolio (study ID: 49429) and is registered on the ISRCTN trial database (ISRCTN87404878). The trial Sponsor was the Newcastle Upon Tyne Hospitals NHS Foundation Trust. The trial was conducted according to the 1964 Declaration of Helsinki principles and its later amendments. All participants provided written informed consent. A summary of the study visits is shown in Supplementary Figure 1.

### Study population

Participants were recruited from primary and secondary care centres in the Northeast of England. Participants were eligible for inclusion if they were aged 65 years and over and had probable sarcopenia according to the European Working Group on Sarcopenia in Older People (EWGSOP) 2019 definition of sarcopenia [15], operationalised as low maximum handgrip strength (<16kg for women, <27kg for men) or a prolonged five times sit to stand time (>15 seconds). Participants also had a 4- metre walk speed of <0.8m/s, which denotes severe sarcopenia in the 2019 EWGSOP guidelines. Low muscle mass was not required for inclusion in the trial; we chose to base trial inclusion on the EWGSOP definition of probable sarcopenia, which does not require low muscle mass and better reflects current clinical practice [16]. The phenotypic frailty score was calculated using the method of Fried et al. [17, 18]

Trial exclusion criteria were selected to (a) avoid enrolment of those with contraindications or at high risk of side effects from Acipimox and (b) exclude participants with skeletal myopathy clearly due to an alternative cause rather than sarcopenia. Eligibility criteria have been published previously in the protocol paper [14] and are shown in Supplementary Table 1.

### Intervention

All participants received Acipimox 250mg capsules [Olbetam; Pfizer Ltd, Sandwich, Kent, UK] orally for three weeks, starting 48 hours after the baseline muscle biopsy and continuing until the day before the second muscle biopsy. The dose was adjusted according to baseline renal function. Participants with creatinine clearance (estimated by the Cockcroft-Gault equation) [19] >60 mL/min were prescribed one capsule three times per day. Participants with creatinine clearance of 45–60 mL/min were prescribed one capsule twice daily. To suppress facial flushing, a commonly reported adverse effect of Acipimox, participants were prescribed aspirin 75 mg orally to be taken 30 min before the first dose of Acipimox each day [20]. Participants who took aspirin as part of their usual medication regime continued their usual dose and were not prescribed additional aspirin.

### Adherence

Participants were asked to bring their unused trial medication to the follow-up visit. Adherence was evaluated by comparing the final tablet count with the initial tablet allocation; adherence was calculated as (the number of tablets taken / the number of tablets expected to be taken by the final study visit) X 100.

### Outcomes

#### Muscle and whole blood biochemistry outcomes

##### Muscle biopsy

Muscle biopsy of vastus lateralis muscle was performed at baseline and follow-up. Biopsies were taken under local anaesthetic, using a Weil-Blakesley conchotome. The second biopsy was taken from the same leg as the first biopsy, a minimum of 3cm from the first biopsy site, to avoid fibrotic or inflammatory changes affecting the results of the second biopsy. Participants were asked to withhold aspirin for 24 hours before the biopsy. Biopsy samples were weighed and then snap-frozen in liquid nitrogen-cooled isopentane immediately after extraction, then stored at −80°C pending analysis.

NAD+/NADH concentrations and ratios were measured using a modified protocol of the Promega NAD/NADH Glo assay kit [Promega, Southampton, UK]. Briefly, frozen muscle samples were homogenised (10mg/ml) in a mixture of equal volumes of PBS and 0.2N NaOH, 1% DTAB at 8,000 rpm (3 x 20s, with 20s intervals, at 0°C) in a Precellys Evolution homogeniser with Cryolys Evolution cooling unit [Bertin Technologie]). Homogenates were centrifuged at 20,000xg for 5 min at 4°C and diluted up to 1:4. NAD+ and NADH were quantified separately from 50µl sample aliquots per the manufacturer’s recommendations, using NAD+ standard curve.

ATP/ADP concentrations and ratio were assessed by luciferase/luciferin chemiluminescence, using a protocol adapted from Porteous et al [21]. Briefly, frozen muscle samples were homogenised (20mg/ml) in perchloric acid solution (3% v/v HCIO4, 2mM Na2EDTA, 0.5% Triton X-100) in a Precellys Evolution homogeniser with Cryolys Evolution cooling unit (Bertin Technologies).

Homogenates were centrifuged at 20,000xg for 5 min at 4°C and supernatants diluted to 1mg/ml. Samples, or ATP and ADP standards prepared in perchloric acid solution, were neutralised with ice-cold 2M KOH, 2mM Na2EDTA, and 50mM MOPS. Precipitates were pelleted by centrifugation (10,000xg for 3 min). For accurate ADP quantification, ATP was removed from aliquots of neutralised extract by adding an equal volume of 100mM Tris/HCl pH 8.0, 10mM MgCl2, 5mM GMP, 20mM Na2MoO4 and 60µg/ml ATP sulfurylase, and incubating at 30°C for 60 min with gentle shaking, followed by 100°C for 5 min. Aliquots (40µl) of neutralised extract (sample and standards) and ATP sulfurylase-treated sample were added to 160µl 100mM Tris/acetate pH 7.75, 2mM Na2EDTA, 50mM MgCl2 in opaque 96-well plates. ADP standards and samples were analysed in duplicate, with one set untreated and one set supplemented with 2.12mM phosphoenolpyruvate and 4.9U/ml pyruvate kinase for 60 min at 25°C, to determine residual ATP. A luciferin/luciferase solution (40µl), consisting of 1.92µg/ml luciferase, 120µM luciferin, 7.5mM dithiothreitol, 0.04% (w/v) BSA, 25% (v/v) glycerol, was added to all samples/standards via an automatic injector in a TriStar LB942 Luminometer (Berthold Technologies). Chemiluminescence was measured and ATP/ADP concentrations quantitated by comparison with appropriate standards.

Cytochrome C oxidase/succinate dehydrogenase (COX SDH) histochemistry [22] and quadruple immunofluorescence [23] were used to assess respiratory chain deficiency. Briefly for COX/SDH staining, COX solution was added to muscle sections for 45min at 37°C, washed and followed by incubation with NBT solution for 45min at 37°C. Sections were washed and dehydrated through graded alcohols before imaging on a Zeiss AxioVision Image Acquisition System where COX-deficient fibres could be identified. Quadruple immunofluorescence was undertaken on 10µm muscle sections as described by Rocha et al [23]. In brief, sections were incubated with primary antibodies, NDUFB8 (complex I), MTCO1 (complex IV), VDAC1 (mitochondrial mass) and laminin (fibre membrane marker), overnight at 4°C. Sections were washed, followed by incubation with secondary antibodies for 2 hours at 4°C and a further incubation with streptavidin conjugated with Alexa Fluor 647nm for 2 hours at 4°C. Following washes, sections were mounted using ProLong gold antifade reagent and imaged using a Zeiss Axioimager microscope. Mitochondrial DNA (mtDNA) copy number was assessed by quantitative PCR by measuring levels of *MT-ND1* (NC_012920.1) and *B2M* (NG_012920.1)[24]. Results are reported as copy number per cell.

### Whole blood

Whole blood was collected and frozen at −80°C. Samples were analysed for NAD+/NADH concentrations and ratios using Q- NADMED Blood NAD+ and NADH assay kit as per the manufacturer’s instructions (NADMED, Helsinki, Finland) to test whether a precision target population (individuals with low skeletal muscle NAD+/NADH ratios at biopsy) could be identified by less-invasive biomarkers.

### Physical performance measures

Physical performance was measured using the Short Physical Performance Battery (SPPB) at baseline and follow-up [25]. Handgrip strength was measured using a Jamar dynamometer (Sammons Preston, Bolingbrook, USA) according to a standard protocol [26]. Three measurements were taken from each side, and the maximum value was used in analyses. Each participant’s real-world mobility was assessed digitally with an Axivity AX6 wearable device Axivity Ltd, Newcastle Upon Tyne, UK; Accelerometer 100Hz, ± 8g, 1 mg (at ± 8g); Gyroscope 100Hz, ≥ ± 2000 dps, 70 mdps (at ± 2000 dps)) that was worn continuously for seven days at baseline and follow-up. The device was attached over the fifth lumbar vertebra (L5) with a medical-grade hydrogel adhesive patch (Nile, Ängelholm, Sweden) and covered with a Hypafix™ bandage. Participants were asked to continue their normal routine. A state-of-the-art analytical pipeline, extensively validated as part of the Mobilise-D project [27, 28] was then applied to the raw wearable device data to calculate clinically relevant walking and gait measures, or digital mobility outcomes (DMOs). In the context of this study, the DMOs of interest were the number of walking bouts undertaken per day, the mean daily walking bout duration (seconds) and the mean walking speed (metres per second) estimated from short walking bouts.

### Magnetic resonance imaging and magnetic resonance spectroscopy outcomes MR spectroscopy

Phosphorus MR spectroscopy (MRS) assessed the phosphocreatine recovery rate (τ_1/2_ PCr) in the calf following exercise. The full MRS analysis protocol has been published previously [29]. Briefly, subjects were requested to perform a low-intensity plantar flexion exercise in the scanner with incremental loading, until phosphocreatine concentrations in gastrocnemius and soleus were depleted by approximately 50%. Measurements were taken every 10s during exercise and for 6 minutes post-exercise. The recovery half-time of phosphocreatine concentration was modelled as a measure of mitochondrial oxidative function, with shorter times implying higher function. Quantitative Dixon MRI was acquired to assess intramuscular fat infiltration in the muscles of the lower limb [30].

### Adverse events coding

Adverse events were collected at each study contact and followed until 28 days after the last participant’s visit. The Chief Investigator and data manager coded events using the Medical Dictionary for Regulatory Activities (MedDRA®) coding system (version 24).

### Sample size

Few data existed on which to base a sample size calculation. The sample size was selected to enable the detection of a one standard deviation change in measures of mitochondrial NAD+/NADH ratios, ATP/ADP ratios, or respiratory chain function. To detect this change with an alpha of 0.05 and 80% power, 11 paired observations are required. We planned to recruit 16 participants to allow for dropouts and non-completion of the medication course.

### Data analysis

The primary outcome of difference in skeletal muscle NAD concentrations between baseline and follow-up was reported as median and interquartile range. The primary analysis was completed as an intention to treat population for all those with data at baseline and follow-up. Related-samples Wilcoxon signed-rank test was used to analyse the primary outcome and continuous non-parametric secondary outcomes, and a paired t-test was used for continuous normally distributed secondary outcomes. Spearman’s Rho was used to test the correlation between nonparametric variables. Further Details are given in the Statistical Analysis Plan provided in the Supplementary Materials. Participants with missing outcome data were excluded from the analysis of that outcome. A two-sided p-value of <0.05 was taken to denote statistical significance for all analyses. No adjustments were performed for multiple testing. Statistical analyses were completed using SPSS Statistics version 29.0.1.1.0 (171) [IBM, New York, USA]

## Results

Between October 2022 and September 2023, 75 participants expressed an interest in participating in the trial. Of these, 24 were potentially eligible and willing to take part after the pre-screening process, and 11 passed screening. The CONSORT diagram (Figure 1) and Supplementary Table 2 show participant flow through the trial. The first 11 participants completed the final outcome visit and took the study medication correctly; therefore, recruitment was stopped as the target number of participants for analysis had been achieved. Details of the study participants at screening are shown in Table 1. All fulfilled the criteria for probable sarcopenia under the EWGSOP2 guidelines. Participants had impaired physical performance, as evidenced by the slow walk speed and prolonged sit-to-stand times. None of the participants discontinued the study medication prematurely. Median medication adherence was 95% (range 91–104%).

**Figure 1.**
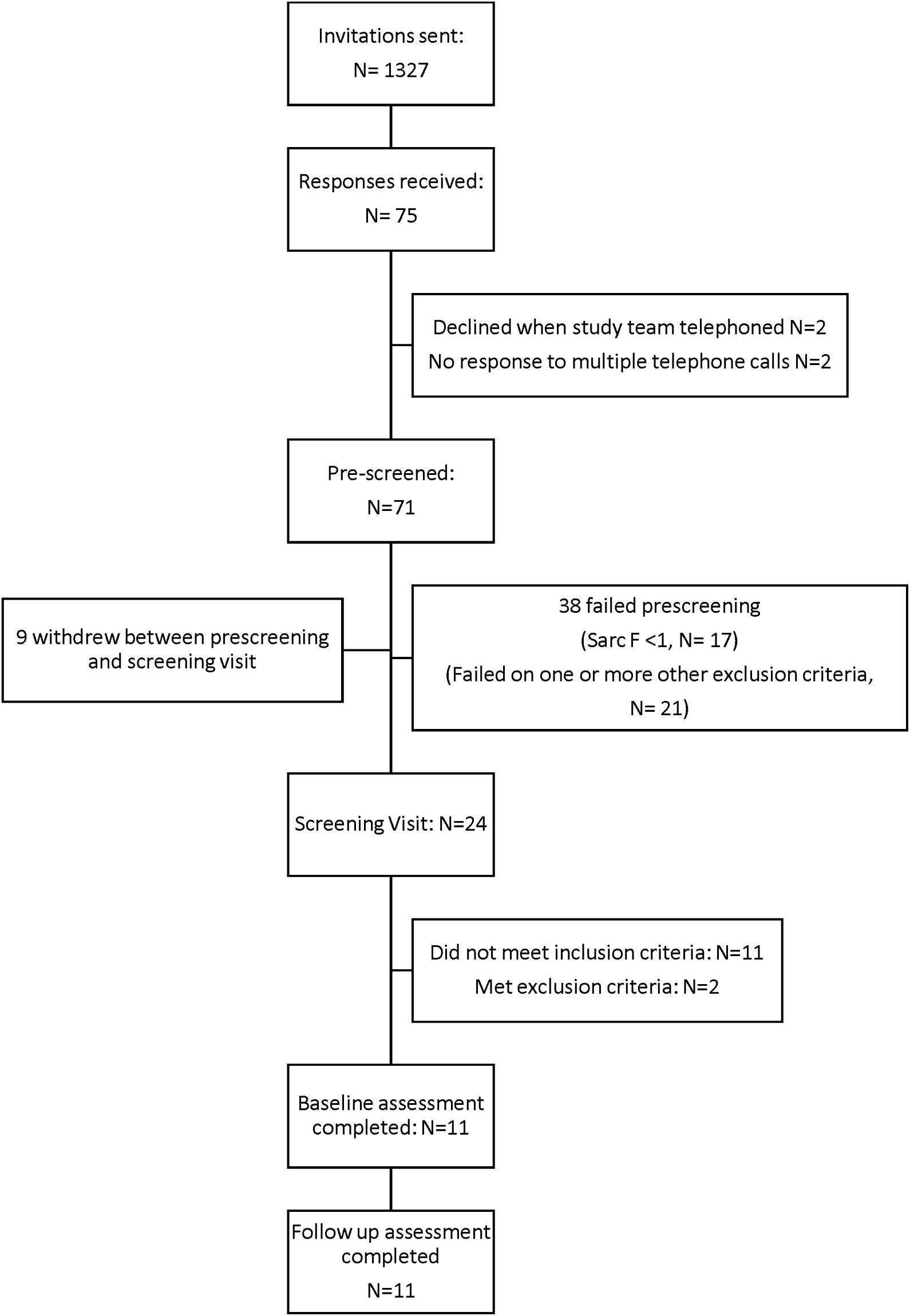
CONSORT Diagram of participant flow through the trial.

**Table 1.** Participant characteristics at screening (n=11)

|  | N (%) |
| --- | --- |
| Mean age (years) (SD) | 78.9 (4.3) |
| Female sex (%) | 8 (73) |
| White British ethnicity (%) | 11 (100) |
| Fried Frailty state<br>• Robust (%)<br>• Pre frail (%)<br>• Frail (%) | 5 (45)<br>5 (45)<br>1 (9) |
| *Comorbidity* |  |
| Cardiovascular or cerebrovascular disease (%) | 0 (0) |
| Diabetes (%) | 0 (0) |
| Atrial Fibrillation (%) | 0 (0) |
| Hypothyroidism (%) | 1 (9) |
| Hyperthyroidism (%) | 0 (0) |
| Anxiety (%) | 2 (18) |
| Depression (%) | 2 (18) |
| Parkinsonism (%) | 0 (0) |
| Chronic obstructive pulmonary disease (%) | 2 (18) |
| Asthma (%) | 0 (0) |
| Osteoarthritis (%) | 7 (64) |
| Mean maximal handgrip strength (men) (kg) (SD) | 33.3 (4.2) |
| Mean maximal handgrip strength (women) (kg) (SD) | 16.5 (2.3) |
| Median 5x sit-to-stand time (s) (range) | 17.9 (16.0-18.5) |
| Mean 4m walk speed (m/s) (SD) | 0.69 (0.07) |
| Mean BMI (kg/m<sup>2</sup>) (SD) | 25.2 (3.5) |
| Creatinine Clearance (ml/min) (SD) | 58.5 (11.6) |

### Muscle and Blood biochemistry

Eight participants were included in the primary analysis (NAD was out of range in samples from 2 participants at baseline and follow-up, and a further 1 participant at follow-up). NAD and NADH concentrations are shown in Table 2. There was no statistically significant difference in the primary outcome of change in NAD concentrations in skeletal muscle between baseline and follow-up [median difference: −0.003 umol/g (IQR −0.06 to 0.02); P=0.26]. Similarly, no statistically significant differences were observed in NADH concentrations or NAD/NADH ratios. Details of the secondary outcomes analysed from skeletal muscle biopsy samples are given in Table 2. There were no statistically significant changes in any of these measures between baseline and follow-up.

**Table 2:**
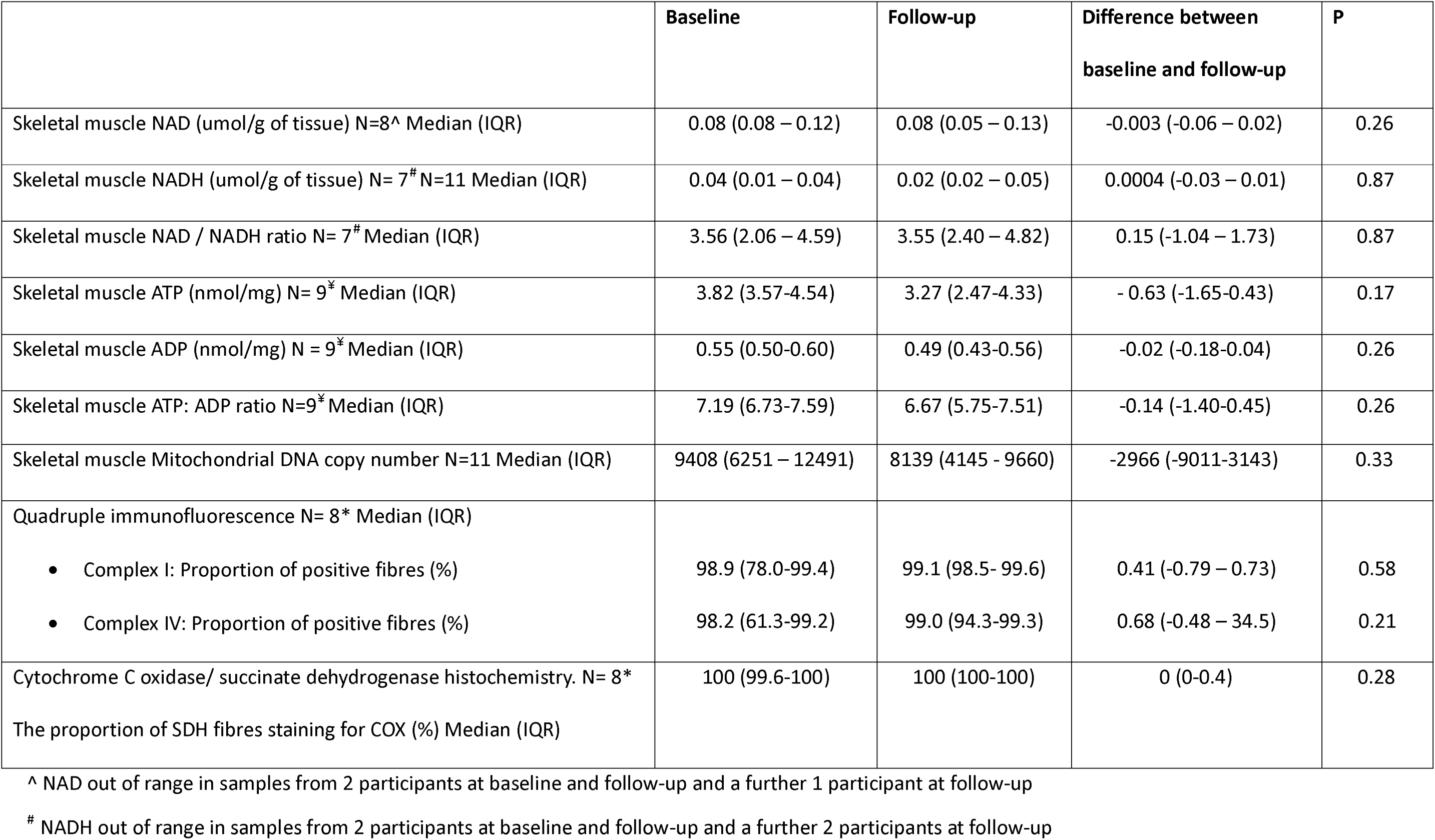
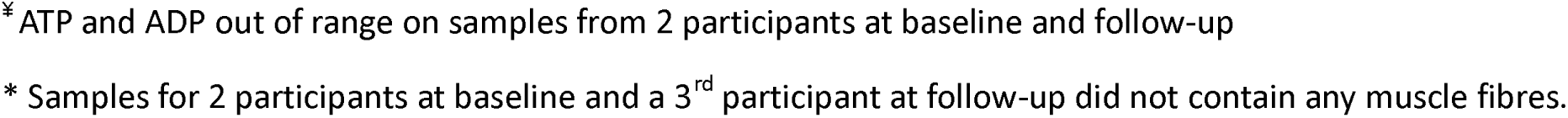
Change in skeletal muscle measures between baseline and follow-up.

Similarly, there was no statistically significant difference in the change in NAD concentrations or NADH concentrations in whole blood [median difference for NAD: 1.02 umol/L (IQR −1.24 to 1.64); P=0.25; median difference for NADH: −0.21umol/L (IQR −0.53 to 0.09); P=0.16]. Spearman’s correlations between whole blood NAD and NADH concentrations and skeletal muscle NAD and NADH concentrations showed inconsistent and highly variable results; only the inverse association between skeletal muscle and whole blood NADH concentrations at baseline approached statistical significance (rho = −0.67; p=0.05). Details are shown in Supplementary Table 3.

### Measures of Physical Performance and MRI outcomes

No statistically significant changes in physical performance measures were recorded during clinic visits or DMOs, Table 3. All subjects successfully completed MR studies at both time points, and all data was analysable. Phosphocreatine recovery time (τ_1/2_ PCr) was longer at follow-up than at baseline; [mean change in τ_1/2_ PCr 7.6 seconds (SD 11.5); P=0.05. MR Spectroscopy results are shown in Table 4.

**Table 3:** Change in strength and physical performance measures between baseline and follow-up.

|  | **N** | **Baseline** | **Follow-up** | **Difference**<br>**between baseline**<br>**and follow-up** | **P** |
| --- | --- | --- | --- | --- | --- |
| Maximum Grip strength (Kg) men [mean (SD)] | 3 | 30.0 (5.29) | 28.0 (3.46) | 0.2 (2.6) | 0.82 |
| Maximum Grip strength (Kg) women [mean (SD)] | 8 | 17.3 (2.82) | 18.3 (2.71) |  |  |
| 4m walk speed (m/s) [mean (SD)] | 11 | 0.88 (0.17) | 0.86 (0.15) | -0.02 (0.12) | 0.67 |
| SPPB Total Score [mean (SD)] | 11 | 8.09 (2.39) | 8.36 (1.50) | 0.27 (1.79) | 0.63 |
| Walking Bouts Per Day (number) [mean (SD)] | 9 | 232 (108) | 256 (88) | 16 (75) | 0.55 |
| Mean daily walking Bout Duration (seconds) [mean (SD)] | 9 | 13.0 (2.70) | 12.7 (1.96) | -2.6 (2.2) | 0.73 |
| Mean walking speed in short bouts (m/s) [mean (SD)] | 9 | 0.66 (0.10) | 0.66 (0.07) | 0.02 (0.04) | 0.21 |

**Table 4.** MR Spectroscopy Measures.

|  | **Baseline** | **Follow up** | **Difference between baseline**<br>**and follow-up** | **P** |
| --- | --- | --- | --- | --- |
| MRI Spectroscopy $\tau_{1/2}$PCr N=11 [Mean (SD)] | 29.5 (6.9) | 37.1 (13.9) | 7.6 (11.5) | 0.05 |
| Fat content of lower limb skeletal muscle (%) [mean (SD)] | 8.9 (2.1) | 8.9 (2.2) | 0 (0.5) | 0.85 |

### Safety and adverse events

Seven participants reported one or more adverse events. In total, 19 AE were reported; 8 were related to a study drug or procedure, and 11 were unrelated. None were reported as serious adverse events. Full details are shown in Supplementary Table 4.

## Discussion

We found no evidence that Acipimox 250mg taken for three weeks improved circulating or skeletal muscle NAD concentrations, mitochondrial function or physical performance in older adults with probable sarcopenia. Acipimox was well tolerated with good adherence to the prescribed regime. This contrasts with previous drug trials in sarcopenia, which have had high dropout rates or study medication discontinuation. [31, 32] Despite the known challenges of recruiting and retaining older people with sarcopenia into clinical trials,[33] we successfully recruited and retained participants with significant physical impairments into a detailed, intensive experimental medicine study. Mechanistic studies in sarcopenia have historically been limited due to concerns that this patient group would be unwilling or unable to tolerate repeated invasive procedures such as skeletal muscle biopsy. [34] We demonstrated that repeated muscle biopsy can be performed safely in this group and that ^31^P MR Spectroscopy and 7-day, body-worn activity monitors were well tolerated by older adults living with probable sarcopenia.

The lack of an effect of Acipimox on skeletal muscle NAD concentrations in this study is in keeping with other recent trials of other NAD precursors in humans [9]. The NAD pool in skeletal muscle is maintained through a continuous process of biosynthesis via the salvage pathway and the Preiss-Handler pathway, on the one hand, and is depleted by sirtuins pathway, cluster of differentiation (CD) 38 enzymes and poly(ADP-ribose) polymerase (PARP) enzymes. Acipimox, a synthetic nicotinic acid analogue, requires a functioning Preiss-Handler pathway to generate NAD. Due to differences in enzymatic expression, not all tissues can convert each precursor to NAD with equal efficacy. Notably, the Preiss-Handler pathway, necessary to metabolise Acipimox, is less prominent in skeletal muscle [35]. Animal models have shown that trigonelline (another NAD precursor metabolised by the Preiss-Handler pathway) leads to a small increase in NAD in the liver, kidney and muscle in young mice but only in the liver and kidney of aged mice [7]. These data raise the possibility that the Preiss-Handler pathway is insufficiently active in older sarcopenic muscle to metabolise Acipimox to NAD. NAD precursors metabolised via the salvage pathway (e.g nicotinamide or nicotinamide riboside) offer an alternative therapeutic approach. Another possible explanation is that age-related increases in the enzymatic consumption of NAD through PARPs and CD38 enzymes might simply swallow up additional NAD generated from supplementation. Accumulations in DNA damage and inflammation as a result of ageing lead to chronic activation of PARPs and CD38 [36] [37] [35]. If the increased enzymatic consumption of NAD is the primary cause of NAD depletion in sarcopenia, any NAD increase from Acipimox may have been outweighed by this consumption, resulting in a failure to boost the NAD pool. Elhassan et al found that although the NAD precursor, Nicotinamide Riboside (NR), failed to increase in skeletal muscle NAD in older men, it did increase other components of the NAD metabolome. [38]

Skeletal muscle NAD concentrations in the current study were higher than those reported in previous studies [6]. We did not require low muscle mass for inclusion, and physical performance measures were better than seen in some previous trials although still within the range sufficient to diagnose probable sarcopenia. If individuals were NAD replete, supplementation is unlikely to increase NAD concentration. The normal range for NAD levels in skeletal muscle in healthy ageing and in sarcopenia is not known and it is not currently feasible to target a specific population that is NAD- deplete; the lack of correlation between NAD concentrations in whole blood and skeletal muscle NAD concentrations in our study suggests that it may not be possible to use blood NAD concentrations to discern the repletion status of NAD in skeletal muscle.

We did not find any evidence to suggest that Acipimox was associated with increased mitochondrial activity. This contrasts with two studies which showed that short-term use of acipimox improved skeletal muscle mitochondrial function. Daniele et al. found that ATP synthesis significantly increased *ex vivo* in the fresh muscle tissue of 11 obese normal glucose tolerant and 11 T2DM patients after 12 days of treatment with acipimox 250mg four times a day. (Daniele, 2014 #87) De Weijer et al. demonstrated increased *ex vivo* mitochondrial respiration in the skeletal muscle of patients with T2DM treated with Acipimox 250mg three times a day for two weeks. (Van De Weijer, 2015 #25) Elevation in the expression of nuclear-encoded mitochondrial gene sets was also observed. Although the authors speculate that the improved mitochondrial function was due to NAD supplementation, no direct evidence is presented to support this hypothesis. In line with our findings, Van De Weijer et al. found no increase in mitochondrial DNA (mDNA).

The results of our study are consistent with a previous study of obese individuals (without T2DM) treated with Acipimox 250mg thrice day for 6 months, which also did not find any impact on mitochondrial function in muscles across various methods, including ^31^P MR spectroscopy [39]. Our MR spectroscopy data showed a marginally significant increased τ_1/2_ PCrbetween baseline and follow-up. This may suggest reduced mitochondrial function; however, both baseline and follow-up findings could be considered normal for a population of this age and as this finding was only marginally significant and the sample size small, this likely reflects a chance finding. It is, however, also possible that Aspirin or Acipimox causes an unexpected increase in PCr recovery time. PCr recovery time increases when substrate delivery to the muscle is reduced (e.g. due to reduced blood flow or glucose uptake) or when mitochondrial function is impaired. Aspirin has been found to reduce reactive hyperaemia in healthy volunteers and in younger and older recreationally active men [40, 41]; Acipimox has been shown to reduce cardiac ejection fraction (and hence cardiac output) in rats but not in humans [42]. Finally, Acipimox acts on the niacin receptor GPR109a expressed on pancreatic beta cells, reducing glucose-induced insulin secretion and cellular glucose uptake. It is unclear whether any of these putative mechanisms contributed to the observed result; further studies would be needed to clarify this issue.

Given that Acipimox did not increase NAD concentration or mitochondrial function in this study, it is unsurprising that Acipimox also failed to improve measures of muscle strength or physical performance. Despite promising preclinical results suggesting NAD supplementation improves muscle function in nematode worms, mice, and rats, human intervention trials of Acipimox and other NAD precursors have not delivered similar improvements. None have improved gait speed, and most have failed to demonstrate improvements in strength, physical activity, or body composition. [10]. It could be argued that the chosen dose or duration of therapy may have been insufficient to produce the desired effect, especially on physical function. However, previous studies have shown biological effects after 12-14 days of treatment at doses similar to those used in the current study (250mg three or four times a day). [43] [12, 39].

### Strengths and Limitations

The key strength of our study was our ability to recruit and retain older people with probable sarcopenia, conducting a detailed experimental medicine protocol in this heretofore challenging population. Patients with sarcopenia are usually prefrail or frail, and many of them have multiple long-term conditions, which makes it challenging to recruit them for clinical trials. [31] Despite the rigorous protocol, the study had exceptional retention rates, with all participants completing the final trial visit and demonstrating over 90% adherence. This indicates that experimental medicine studies in sarcopenia are feasible.

Several limitations should also be acknowledged. The study population was recruited from a single centre in the Northeast of England, a region with limited ethnic diversity, potentially limiting the generalisability of these findings. Furthermore, the study population is not fully representative of the population living with sarcopenia. Efforts were made to minimise the study exclusion criteria. However, Acipimox interacts with HMG-CoA reductase inhibitors (statins) and muscle biopsy is contraindicated in people prescribed anticoagulant or anti-platelet therapy other than aspirin. In the UK population aged between 40 and 99 years, 25% are prescribed a statin, 12% are prescribed an antiplatelet agent, and 4% are prescribed an anticoagulant. [44] Rates of prescribing of these medications increase with increasing age in the UK [44–46] It is, therefore, likely that some key groups (particularly those with metabolic syndromes, vascular disease and dementia) were underrepresented in this study.

The small sample size of our study raises the possibility of a type 2 error. Eleven paired observations (using a paired t-test) were required to detect a change with an alpha of 0.05 and 80% power. We had originally planned to recruit 16 participants to allow for dropouts and non-completion of the medication course. However, the first 11 participants completed the follow-up assessment and continuing to recruit would have placed further participants at risk from biopsy and study medication side effects without justifiable scientific gain. While muscle biopsies were performed on all 11 participants at baseline and follow-up, not all samples were suitable for analysis. Despite appearing to be muscle when biopsied, some samples had extensive fatty infiltration or fibrous tissue, making them unsuitable for analysis. Muscle atrophy is a known factor in failed biopsies. In one group, ultrasound guidance showed that only 18.7% of frail, older women had ideal muscle thickness for biopsy, while 68.8% had sub-optimal thickness and 12.5% had inadequate muscle. [34]. While ultrasound is not typically used to choose biopsy sites, it could have helped identify areas with better muscle thickness, leading to improved samples. In future studies, sample size calculations and recruitment targets should consider the difficulties of obtaining enough muscle tissue in this population.

### Future work

The consistent failure to replicate pre-clinical findings with NAD precursors in humans suggests that NAD metabolism in humans, especially in older individuals and those with sarcopenia, is insufficiently understood and may differ in important ways from other species. Future studies should focus on understanding the factors causing NAD depletion in sarcopenia. Inhibiting NAD consumption, alone or in combination with NAD precursors, may provide a more productive approach to enhancing the NAD pool. Studies in mice have shown that manipulating levels of PARP-1 and CD38 can increase NAD levels, boost sirtuins, boost mitochondrial gene expression, and preserve muscle function during aging. [35, 47, 48].

PARP and CD 38 inhibitors are now licensed in the USA and Europe for therapeutic use in cancer. However, current CD 38 and anti-PARP antibodies are cytotoxic. [49] Anti-CD38 antibodies that specifically inhibit CD38 NADase activity without cytotoxic effects could become an important tool for enhancing NAD in age-related diseases. Experimental medicine studies similar to the one that we report here will be crucial in evaluating these new therapies.

## Data Availability

All data produced in the present study are available upon reasonable request to the authors

## Competing interests and funding

### Funding

- MRC Confidence in Concept grant
- CM, MDW, AAS, GG, LR acknowledge support from the NIHR Newcastle BRC

### Conflicts of interest

MDW: Rejunvenate BioMed consultancy

## Notes

### Clinical Trial

ISRCTN trial database (ISRCTN87404878)

### Clinical Protocols

https://bmjopen.bmj.com/content/14/2/e076518.abstract

### Author Declarations

The trial was approved by the UK Health Research Authority Northeast - Tyne & Wear South Research Ethics Committee (approval number 21/NE/0100). The trial was also approved by the UK Medicines and Healthcare Products Regulatory Agency (MHRA; EudraCT trial reference number 2021-000993-28). The trial has been included in the National Institute for Health Research Clinical Research Network (NIHR CRN) portfolio (study ID: 49429) and is registered on the ISRCTN trial database (ISRCTN87404878). The trial Sponsor was the Newcastle Upon Tyne Hospitals NHS Foundation Trust. The trial was conducted according to the 1964 Declaration of Helsinki principles and its later amendments. All participants provided written informed consent

