## Supplemental tables for "Effect of Acipimox on skeletal muscle biochemistry, structure and function in older people with probable sarcopenia: an experimental medicine study"

### **Supplementary Data**

**Supplementary Table 1: Eligibility Criteria**

**Eligibility Criteria**

*Inclusion Criteria*

ALL of the following inclusion criteria must be met:

-	Age 65 years or over

-	Low maximum handgrip strength (<16kg for women, <27kg for men) OR five-times sit-to-stand time >15 seconds (inability to complete sit-to-stand test will count as time > 15 seconds)

-	Walk speed <=0.8 m/s on 4 metre walk test

*Exclusion Criteria*

General:

-	Allergy to Acipimox or other niacin-related products

-	Allergy or intolerance of aspirin

-	Any contraindication to taking aspirin

-	Unable to give written informed consent

-	Currently enrolled in another intervention study (observational studies are permitted)

-	Currently participating in supervised exercise classes or physiotherapy

-	Any progressive neurological or malignant condition with a life expectancy <6 months

Safety of IMP and nIMP:

-	Creatinine Clearance <45ml/min (by Cockcroft-Gault equation)

-	Taking statin medication or fibrate medication

-	Active peptic ulcer disease or dyspepsia

Safety of muscle biopsy and MRI:

-	Platelets <100x109/L at screening (contraindication to muscle biopsy)

-	Presence of a bleeding diathesis or use of oral or parenteral anticoagulant medication

-	Antiplatelet agents other than low dose (75mg once daily) aspirin

-	Contraindications to MRI scanning (mild claustrophobia is not a contraindication)

-	Allergy to local anaesthetic (lidocaine)

-	Unable to palpate vastus lateralis muscle to enable biopsy localisation

Other causes of skeletal myopathy:

-	Liver function tests (bilirubin, ALT, alkaline phosphatase) > 3x ULN

-	Symptomatic (NYHA class II-IV) chronic heart failure (diagnosed according to European Society of Cardiology guidelines)

-	Severe COPD (GOLD stage IV)

-	Known myositis or other established myopathy

-	Self-reported weight loss of >10% in the last six months (to exclude significant cachexia)

-	Known uncontrolled thyrotoxicosis

-	7.5mg/day or greater prednisolone use (or equivalent)

**Supplementary Table 2: Reported Exclusion criteria during the screening process.**

**Reported Exclusion Criteria (some participants reported more than one exclusion criterion)**

- Progressive neurological or malignant condition with life expectancy <6 months (N=2)
- Symptomatic (NYHA class II-IV) chronic heart failure (N=5)
- Severe COPD (GOLD stage IV) (N=1)
- 10% weight loss (N=5)
- Taking statin medication or fibrate medication (N=9)
- Use of oral or parenteral anticoagulant medication (N=5)
- Antiplatelet agents other than low-dose aspirin (N=2)
- Allergy to aspirin (N=1)
- Contraindications to MRI scanning (N=2)
- Corticosteroid equivalent to >7.5mg oral prednisolone (N=2)

**Supplementary Table 3: Spearman's correlations between blood NAD / NADH and muscle NAD / NADH.**

| **Baseline** | Rho (p) |
| --- | --- |
| Whole blood NAD vs Skeletal muscle NAD | -0.08 (p=0.83) |
| Whole blood NADH vs Skeletal muscle NADH | -0.67 (p=0.05) |
| Whole blood NAD/NADH ratio vs Skeletal muscle NAD/NADH ratio | -0.50 (p=0.17) |
| **Follow up** |  |
| Whole blood NAD vs Skeletal muscle NAD | 0.19 (p=0.65) |
| Whole blood NADH vs Skeletal muscle NADH | -0.57 (p=0.18) |
| Whole blood NAD/NADH ratio vs Skeletal muscle NAD/NADH ratio | 0.07 (P=0.88) |

**Supplementary Table 4: Summary of adverse event frequencies**

|  | | Number of participants (N=11) |
| --- | --- | --- |
| Number of participants with at least one AE | | 9 |
| Number of participants with at least one AR | | 5 |
| Number of participants with at least one SAE | | 0 |
| Number of participants with at least one SAR | | 0 |
| Number of participants with at least one SUSAR | | 0 |
| Total number of AEs | | 19 |
| Number of the most common AEs: | Itch | 1 |
|  | Soft tissue injury to the leg | 1 |
|  | Flushing | 1 |
|  | Cellulitis | 2 |
|  | Covid 19 Infection | 1 |
|  | Hematoma | 2 |
|  | Aching legs | 1 |
|  | Rash | 1 |
|  | Bruising to arm | 1 |
|  | Sciatica | 1 |
|  | Fall | 1 |
|  | Constipation | 1 |
|  | White coat hypertension | 1 |
| Number of hospital admissions | | 0 |
| Number of deaths | | 0 |

AE: Adverse even
